# Deconditioning in people living with dementia during the COVID-19 pandemic: findings from the Promoting Activity, Independence and Stability in Early Dementia (PrAISED) process evaluation

**DOI:** 10.1101/2020.11.16.20231100

**Authors:** Claudio Di Lorito, Tahir Masud, John Gladman, Maureen Godfrey, Marianne Dunlop, Rowan H. Harwood

**Affiliations:** Division of Rehabilitation, Ageing and Wellbeing, University of Nottingham, Nottingham NG7 2TU, United Kingdom; Nottingham University Hospitals NHS Trust; Division of Health Sciences, University of Nottingham, Nottingham NG7 2TU, United Kingdom

**Keywords:** deconditioning, physical activity, exercise, dementia, COVID-19

## Abstract

**Background:** Restrictions introduced in response to the COVID-19 pandemic led to increased risk of deconditioning in the general population. No empirical evidence of this effect however has been empirically gathered in people living with dementia.

**Objective:** This study aims to identify the causes and effects of COVID-19-related deconditioning in people living with dementia.

**Design:** Longitudinal phenomenological qualitative study.

**Subjects:** Participants living with dementia, their carers and therapists involved in the Promoting Activity, Independence and Stability in Early Dementia (PrAISED) process evaluation during the COVID-19 pandemic.

**Methods:** Qualitative interviews with participants were conducted remotely at two time points. The data were analysed through deductive thematic analysis.

**Results:** Twenty-four participants living with dementia, 19 carers and 15 therapists took part in the study. A self-reinforcing pattern was common, whereby lockdown made the person apathetic, demotivated, socially-disengaged, and frailer. This reduced activity levels, which in turn reinforced the effects of deconditioning over time. Without external supporters, most participants lacked the motivation / cognitive abilities to keep active. Provided the proper infrastructure and support, some participants could use tele-rehabilitation to combat deconditioning.

**Conclusion:** The added risks and effects of deconditioning on people with dementia require considerable efforts from policy makers and clinicians to ensure that they initiate and maintain physical activity in prolonged periods of social distancing. Delivering rehabilitation in the same way as before the pandemic might not be feasible or sustainable and innovative approaches must be found. Digital support for this population has shown promising results, but still remains a challenge.

## Introduction

Sir Muir Gray and Dr William Bird in an opinion letter published by the BMJ Opinion as the first wave of COVID-19 in the UK was subsiding [1] stated that*”Months of isolation and reduced levels of activity at home will have an immense deconditioning effect on millions of people”*. A survey carried out in the UK found that 30% of adults aged 50+ reduced activity levels in the first wave of the COVID-19. About 30% of respondents reported they lacked motivation to exercise [2].

Compared to the normative population, individuals living with dementia face added challenges to being physically active, including impairment of cognitive and motor skills [3-5] and greater apathy or lack of motivation to get active and maintain activity levels [6]. This population is therefore even more exposed to the risks of de-conditioning. The risk of deconditioning in people living with dementia was further compounded by governments’ shielding advice for vulnerable people to decrease the risk of contracting the virus [7], thus reducing opportunities to do physical activity outdoors.

The benefits of physical activity and exercise for people living with dementia have long been established. These include the maintenance of physical abilities, independence, and quality of life over time [8]. There is also evidence around the effects of sedentariness, including loss of balance, increased risk of falls and mental ill-health [9]. It has been found that even a few days of sedentary lifestyle generate muscle loss, neuromuscular junction damage and fibre denervation, insulin resistance, decreased aerobic capacity, fat deposition and low-grade systemic inflammation [10]. No empirical evidence on the effect of COVID-19 pandemic in a population living with dementia is available.

A unique opportunity to study this phenomenon was presented in the context of the Promoting Activity, Independence and Stability in Early Dementia (PrAISED) randomised controlled trial (RCT) [11]. PrAISED is evaluating an individually-tailored physical activity and exercise intervention. Mode of delivery was changed during pandemic lockdown from face-to-face to remote video- or telephone support, where possible. Alongside the RCT, we performed a process evaluation using qualitative interviews [12] to gather data on the impact of the pandemic on physical activity and overall wellbeing.

With the aim of improving the ability to reduce the effect of restrictions to activity in people with dementia and how they might be overcome, we used this opportunity to collect and analyse empirical data from the PrAISED process evaluation to answer two research questions:

1. What were the causes of deconditioning for people living with dementia during the COVID-19 pandemic?
2. What were the effects of deconditioning for people living with dementia and their caregivers during the COVID-19 pandemic?

## Methods

This study was grounded in a phenomenological qualitative approach, making use of data triangulated from interviews with participants living with dementia, their carers and the therapists involved in the PrAISED process evaluation during the COVID-19 pandemic. The PrAISED trial and process evaluation have received ethical approval number 18/YH/0059. The ISRCTN Registration Number for PrAISED is 15320670.

### Sample

The main researcher (CDL) accessed the PrAISED RCT database - for selection criteria of participants in the RCT see: [11] - and purposively selected participants to ensure an equal representation of geographical location (Nottinghamshire, Derbyshire, Lincolnshire, Somerset and Oxfordshire), gender, residence status (living with carer or independently), and RCT arm (i.e. intervention or control group). The carers of the selected participants who agreed to take part in the study were also invited to take part in the interviews. No participant living with dementia or carer refused to take part. All the therapists involved in the delivery of PrAISED during the COVID-19 pandemic (n=16) were contacted by the main researcher and invited to take part in the study. Only one refused.

### Data collection

Interviews were conducted at two time points, the first set two months after the national lockdown was imposed in England (i.e. May 2020), the follow up two months after the first set (i.e. July 2020). The interviews were conducted by a researcher (CDL) remotely, either by phone (i.e. a speaker phone for participants and carers to be able to contribute) or video conferencing, depending on the participants’ and therapists’ preference. Prior to the interview session, CDL mailed (or emailed) a copy of the information sheet and consent form. Verbal consent was recorded on tape on the interview day, before the interview started.

The interviews were guided by a topic guide exploring the impact of the lockdown on exercise, activity and mental well-being. The topic was developed in collaboration with two PPI contributors (MG and MD), who helped to make the questions relevant, meaningful and accessible for the participants. The PPI contributors were also included as co-authors in this paper. The topic guide was used flexibly, to ensure that new topics emerging during the sessions were explored. The interviews were all digitally audio-recorded on a password protected device approved by governance. Sample size was not defined a priori and interviews were carried out until conceptual density (i.e. a *sufficient depth* of understanding of the topic under investigation) [13] was reached.

### Data analysis

The data were analysed through deductive thematic analysis [14]. Two main themes were established a priori, based on the study objectives: 1. Causes of deconditioning as a result of the COVID-19 pandemic; 2. Consequences of deconditioning as a result of the COVID-19 pandemic. A transcription agency transcribed the interviews verbatim. The transcripts were passed to CDL, who fully anonymised them. CDL then familiarised with the transcripts and noted down preliminary analytical notes.

Coding of a sample of three transcripts was then carried out by CDL and a second researcher within the research team (TM), who extrapolated segments of the transcripts and categorised them into the themes, independent of each other. The two researchers convened to compare their coding and resolve any potential disagreement. CDL then coded the whole set of transcripts in NVivo 12 [15]. Results of the analysis were fed back to the PPI contributors, who provided their comments on the relevance of findings.

## Results

Twenty four participants living with dementia and 19 carers (five participants wished to be interviewed alone) took part in this study (Table 1). Participants’ mean age was 81 (range: 67-95, SD = 7). Fourteen participants (58%) were male and 19 (80%) lived with the carer. Carers’ mean age was 73 (range: 50-85; SD = 9). Twenty-three interviews (96%) were done through speaker phones and one (4%) through webcam.

Fifteen therapists delivering PrAISED during lockdown were interviewed (Table 2). Four services (80%) delivering PrAISED were represented (i.e. Nottinghamshire, Derbyshire, Lincolnshire, Oxfordshire), but no therapist from Somerset was available for the study. Six therapists (40%) were occupational therapists, five (33%) rehabilitation support workers, and four (27%) physiotherapists. All therapists but one (93%) were female. All interviews were done through the phone.

To keep the participants’ identity confidential, quotes below are reported anonymously. For information on participants’ characteristics for each quote (i.e. gender and age group), please contact the main author.

### Theme 1: Causes of deconditioning in people living with dementia during the COVID-19 pandemic

Individual-specific socio-demographics and certain environmental, intra and inter-personal variables mediated activity levels. Health issues including physical frailty, stage of dementia, pre-existing psychological morbidity (e.g. depression, anxiety), a history of previous injuries and hospitalisation were all linked to a reduced level of physical activity. This was often compounded by reduced access to health care services during the pandemic:

*“I have had a problem with the left hand leg and now the right hand leg, and that’s throwing me out completely. I haven’t been able to get in touch with somebody who could actually help because of lockdown”*. Participant with dementia.

A lack of internal motivation to exercise was quite common and several participants who lived independently observed that because the therapists were not visiting, they could not rely on the external pressure to exercise. On the contrary, pressure from carers helped establish a daily routine of exercise, which was instead linked to prevention of deconditioning:

*I used to do them with an instructor but now I skip the odd ones and I know I shouldn’t be skipping them because it is important. But somebody coming round to knock on the door to take you through them, you just do it”*. Participant with dementia.

*“We (carer and participant living with dementia) always have a walk once a day, so it is a routine. Routines are very rigid and this gives me reassurance that L. is going to do it”*. Carer of participant with dementia.

Another key factor affecting exercise levels was the ability of participants to access and use digital support. Traditional means of communication (phone) did not prove effective in supporting participants. Because of age and / or dementia, most participants had difficulties hearing the therapists or communicating. Thus, phone communications ended up occurring only between the therapists and carers:

*“Supporting participants through the phone is limited. Over the phone it’s difficult to understand if the participant is hearing or acknowledging what I am saying because you can’t rely on non-verbal cues. So you end up speaking with the carer, which I find totally disempowering for the participant”*. Physiotherapist.

As a result, the participants who lacked the digital infrastructure or literacy to be able to keep (visual) social contacts with the therapists reported greater apathy and lower motivation to keep active:

*“It is the face to face interaction I am missing most. And I haven’t got any enthusiasm to do anything that I would normally do. I am sitting in my garden listening to the birds watching the days go by”*. Participant with dementia.

The situation was reversed for participants who felt confident enough to use IT to receive support from the PrAISED therapists through video-consultations. In these circumstances, the social opportunity, support and motivation provided by a professional “outside the family circle” was instrumental for intervention uptake and adherence:

*“Mom just likes to be able to see someone’s face, someone different, she finds it easier to understand the exercises they show her, as opposed to remind her and explain how to do something. Seeing her (the therapist) encourages mom so much!”* Carer of participant with dementia.

The greater need for emotional support among participants was such that the PrAISED therapists reported a paradigm shift in PrAISED delivery, from pre-COVID sessions mostly focused on doing the exercises to a more holistic focus on emotional counselling:

*“It (PrAISED) has actually turned more into a general wellbeing chat, trying to give them positive re-enforcement that they are doing a good job and if they are struggling then encourage them to open up”* Occupational Therapist.

Participants who lived independently (with no in-house informal carers) and were in the control group in PrAISED (thus receiving no regular support and contact) were most exposed to the risk of becoming inactive. This suggests that positive encouragement can prevent deconditioning. Another key environmental factor affecting physical activity levels was the availability of a private outdoor area (e.g. garden). Because walking and gardening were the most frequently reported physical activities, having a garden, as opposed to not having a private outdoor space, helped participants keep physically active:

*“We (carer and participant living with dementia) have got a fair sized garden, so myself, I do a lot of gardening which I really enjoy. And I also like sitting and admiring. That really lifts me up”*. Participant with dementia.

One participant reported how staying indoors had greatly limited her chances of doing physical activity, reinforcing the evidence that during the COVID-19 pandemic, socio-economic status might be linked to a greater risk of deconditioning and ill health:

*“When this first started out I was walking round our block of flats to keep me supple a bit. But then I got a letter that told me not to go out. I live in blocks of flats and I’m on the downstairs one so I only have a tiny little patio and cannot walk in there”*. Participant with dementia.

In terms of environmental factors, the cancellation of activities in the community drastically reduced the opportunities available for participant to keep active:

*“Lots of place have shut down because of COVID, so all the trips and the outings, I mean we used to do Nordic walking as a group. Now, even if he (participant) wanted to keep doing some things, obviously there is no way”*. Carer of participant with dementia.

It was observed that deconditioning risk also greatly depended on intra and inter-personal resources to adapt to the changing circumstances. A participant who was carrying out interviews with community dwellers to build a family tree reported he was now doing research online. Another participant, who used to go to the pub for Quiz Friday reported that his family was organising weekly phone quizzes during lockdown. These new exciting projects had prevented a lack of motivation and apathy to keep active:

*“I like to keep things in order, and because I was very busy before (lockdown), I was neglecting the garden. So I decided right, I am going to concentrate on my garden now, so that is what I have been doing”*. Participant with dementia.

### Theme 2: Effects of deconditioning in people living with dementia during the COVID-19 pandemic

Although to various degrees, deconditioning produced a negative “domino effect”, significantly reverberating on the participants’ physical ability, mental wellbeing, affect, motivation to be socially engaged and their overall quality of life:

*“Because I’m in all the time, I don’t do much walking. It makes me very low and so when I go to bed at night I’m thinking about things and everything’s going through my head and instead of one and one making two, one and one make a hundred. My anxiety is out of control”*.Participant with dementia.

It was observed that the impossibility to progress toward physical health goals that required training outside the home (e.g. swimming pools) had made participants apathetic and demotivated to keep active:

*“The initial goals that the participants had set for themselves during lockdown disappeared from the horizon. So, if a participant wanted to start swimming again, during lockdown this was not even a remote possibility. Therefore, it was difficult for them to keep motivated”*.Rehabilitation Support Worker.

Several participants felt that because of a lack or reduction of exercise, they had lost the progress they had previously achieved. Some carers agreed that dementia symptoms, such as loss of balance and motor skills had visibly progressed. This was particularly noticeable among the participants who were previously supported face-to-face by the PrAISED therapists:

*“I think the things he was doing (dual task exercises with PrAISED therapist) were keeping his brain and body active. I think he has got a bit slower in his walking and speech now”*. Carer of participant with dementia.

This regression made participants lose confidence, resulting in, as eloquently put by one participant, a “lockdown syndrome”, a reluctance and anxiety to leave the home and engage in physical exercise outdoors:

*“The only physical thing that I have been doing lately is walking up and down the garden. I have lost confidence in myself and I need that confidence. I keep saying I should pluck up the courage to go outside of the box that I live in, but I have still not able to get over this locked in syndrome. I tell myself it is what I need to do bit by bit and I am fine with the idea until it comes to actually going”*. Participant with dementia.

The lockdown syndrome was often compounded by gatekeeping behaviour of carers, who were reluctant to encourage the person to go out for fear that they might contract the virus:

*“I still go for a half hour walk around the nearby park everyday but P. (participant living with dementia) has not been with me. I think he would find it difficult to remember to keep the two meter rule. I think it will take me a long time to become confident enough to let him out”*. Carer of participant with dementia.

As a result of some participants becoming more sedentary, carers observed a progression of dementia symptoms and an acceleration of the person’s deterioration. The person’s increased dependency needs and frailty increased the carers’ strain and made them anxious about the person’s health:

*“Because mom is dependent on me for shopping, I mostly come every day. This has bought a level of anxiety that wasn’t there before because she’s got quiet frail and I worry I might bring the infection into the house”*. Carer of participant with dementia.

## Discussion

This qualitative study generated empirical evidence on the causes and effects of deconditioning during the COVID-19 pandemic in a sample of participants living with dementia taking part in the PrAISED process evaluation. The study found that the COVID-19 pandemic (and the consequent lockdown) drastically reduced activity levels in the participants living with dementia taking part in PrAISED, both in the intervention group being supported remotely (compared to the in-person support pre-lockdown) and even more so in control participants, who did not receive any support from PrAISED therapists.

Although deconditioning as a result of social isolation has been reported in the general population and in other risk groups, this study found that having dementia exacerbated the negative effects of deconditioning. Apathy, proneness to anxiety and digital illiteracy all contributed to an added risk of becoming inactive. However, there were some particular issues unique to dementia. The cognitive demands to maintain social distancing rules was a limiting factor likely to be unique to dementia. The risk that the person with dementia might forget social distancing rules (e.g. forgetting to wear a face mask, to keep social distance, to not shake hands, to wash hands) often made carers prone to imposing restrictions to reduce such breaches. However, the imposition of restrictions had negative effects as it limited the opportunities for physical activity (outdoors).

In line with the evidence emerging in the literature [16, 17], it was observed that a self-reinforcing vicious cycle was common among the participants, whereby lockdown made the person apathetic, demotivated, socially-disengaged, frailer and less confident. This markedly reduced activity levels, which in turn reinforced the effects of deconditioning over time. Without external supporters, most participants living with dementia lacked the motivation / cognitive abilities to keep active. In line with research from the Alzheimer Society of Ireland [17], it also highlighted how minimal but consistent input from therapists drastically reduced the risk and effects of deconditioning. Fully embracing the “use it or lose it” philosophy, most participants in the intervention group acknowledged that “some support is better than no support” and affirmed how helpful having a regular contact with the therapist had been throughout the COVID-19 lockdown.

This study is characterised by certain strengths and limitations. It investigated the phenomenon of deconditioning in real time through a rigorous empirical methodology. Further, it collected data at two time points, to monitor progress of deconditioning over time. It presented a timely contribution to current discourse around the phenomenon of deconditioning by presenting the views of people with lived experience of dementia and their caregivers, promoting representation of their perspectives in research [18]. Further, it introduced the views of service providers too, to add a clinical perspective dimension to the data. Because the study was embedded in an existing trial, its sample of participants might not reflect the wider population of people living with dementia. However, the intention of this study was only to report a real time snapshot of the phenomenon in a specific context, to raise awareness and contribute empirical evidence to the existing discourse on the challenges of people living with dementia during the COVID-19 pandemic [19].

Looking forward, this study warrants important reflections for clinical practice and policy. The Physiological Society and Centre for Ageing Better presented recommendations from leading scientists and clinicians in the fields of physiology, nutrition and physiotherapy [2] on how to support older people to remain active and healthy through the pandemic. While the problems identified in this paper have been recognised in the report, the solutions have not been grounded in the experience of people living with dementia. This study illuminates the overlooked but very important issue of people with dementia and provides a grounded evidence based approach to overcoming the challenges faced by this population.

The added risks and consequences of deconditioning on the population of people living with dementia (compared to the general population) requires considerable efforts to be made from policy makers and clinical practitioners. The National Institute for Health and Care Excellence (NICE) states that *‘there is overwhelming evidence that changing people’s health-related behaviour can have a major impact on some of the largest causes of mortality and morbidity’* [20]. It is therefore a health priority to ensure that people living with dementia initiate and maintain exercise and physical activity in current (or future) circumstances requiring prolonged periods of social distancing.

As advocated in a recently published commentary on rehabilitation needs of older people as a result of the COVID-19 pandemic [21], delivering rehabilitation in the same way as before the pandemic will not be feasible, nor practical or sustainable. Innovative ways and approaches must be found. A recent study looking at the effects of deconditioning reported that even low to medium-intensity resistance exercise, easily implementable at home, have positive effects in preserving neuromuscular, metabolic and cardiovascular health [10]. The key then, is to promote motivation and engagement to physical activity in the absence of home-visits from trained professionals.

Our study adds to the mounting evidence [17] that maintaining consistent face-to-face (remote) contact between professionals and people living with dementia (and carers) is an effective strategy to keeping them motivated and active, compared to phone support (or no support at all). While tele rehabilitation has long been established as a successful tool in areas such as pulmonary [22] and stroke rehabilitation [23], there has been limited implementation as a strategy against deconditioning in dementia care. This study found that people living with dementia still experience challenges to accessing digital tele-rehabilitation, given, among other factors, a lack of digital infrastructure, IT literacy and cognitive impairment. Similar findings have been echoed in a recent Dutch study highlighting barriers to the uptake of communication technologies in nursing homes during the COVID-19 pandemic [24].

However, in line with similar attempts to introduce digital technologies in supporting people living with dementia during the COVID-19 pandemic [25, 26], this study showed that provided the proper infrastructure (e.g. digital hardware and software, live support for users to navigate the system, partnership with carers) and balanced integration with the (still preferred) face-to-face interaction [27], in times of need, people living with dementia are willing to use technology [28] to prevent the risks and effects of deconditioning. Investment in digital connectivity, as well as in providing support to enable this population to use tele-rehabilitation in must therefore be scaled up, to ensure readiness for future challenges.

## Data Availability

The data are available from the main author upon reasonable request.

---

Table 1. Characteristics of participants with dementia and caregivers. The table contains information that can potentially make participants identifiable. Readers can contact the corresponding author to request access.

Table 2. Characteristics of therapists and interview sessions. The table contains information that can potentially make therapists identifiable. Readers can contact the corresponding author to request access.

## References

[1] Gray, M, Bird, W. Covid-19 will be followed by a deconditioning pandemic. BMJopinion. https://blogs.bmj.com/bmj/2020/06/15/covid-19-will-be-followed-by-a-deconditioning-pandemic/ (01 November 2020, last accessed)

[2] The Physiological Society and Centre for Ageing Better. Leading scientists call for a National Covid-19 Resilience Programme to keep older people healthy and resilient during lockdown. https://static.physoc.org/app/uploads/2020/11/09152548/A-National-Covid-19-Resilience-Programme-report-web-version.pdf (09 November 2020, last accessed)

[3] Martyr, A, Clare, L. Executive function and activities of daily living in Alzheimer’s disease: a correlational meta-analysis. Dem Ger Cog Dis 2012; 33: 189–203

[4] Giebel, CM, Sutcliffe, C, Stolt, M, et al. Deterioration of basic activities of daily living and their impact on quality of life across different cognitive stages of dementia: a European study. Int Psychoger 2014; 26: 1283

[5] Alzheimer’s Research UK. About dementia. https://www.alzheimersresearchuk.org/about-dementia/?gclid=Cj0KCQjw3JXtBRC8ARIsAEBHg4myuYeahFMLCcSGLr-flQxznmdB-dObW2gXc5MUN9o_dfNw5wwI5EwaAvH6EALw_wcB (26 March 2020, xlast accessed)

[6] Cipriani, G, Lucetti, C, Danti, S, Nuti, A. Apathy and dementia. Nosology, assessment and management. Jour Nerv Ment Dis 2014; 10: 718–24

[7] Public Health England. Guidance on social distancing for everyone in the UK. https://www.gov.uk/government/publications/covid-19-guidance-on-social-distancing-and-for-vulnerable-people/guidance-on-social-distancing-for-everyone-in-the-uk-and-protecting-older-people-and-vulnerable-adults (24 March 2020, last accessed)

[8] Lam, FM, Huang, MZ, Liao, LR, Chung, RC, Kwok, TC, Pang, MY. Physical exercise improves strength, balance, mobility, and endurance in people with cognitive impairment and dementia: a systematic review. Jour Phys 2018; 64: 4–15

[9] Copeland, JL, Ashe, MC, Biddle, SJ et al. Sedentary time in older adults: a critical review of measurement, associations with health, and interventions. Br J Sports Med 2017; 51: 1539

[10] Narici, M, De Vito, G, Franchi, M et al. Impact of sedentarism due to the COVID-19 home confinement on neuromuscular, cardiovascular and metabolic health: Physiological and pathophysiological implications and recommendations for physical and nutritional countermeasures. Eur Jour Sport Sc. 2020; 1–22

[11] Bajwa, RK, Goldberg, SE, Van der Wardt, V et al. A randomised controlled trial of an exercise intervention promoting activity, independence and stability in older adults with mild cognitive impairment and early dementia (PrAISED)-A Protocol. Trials 2019; 20: 1–1

[12] Di Lorito, C, Bosco, A, Goldberg, SE et al. Protocol for the process evaluation of the Promoting Activity, Independence and Stability in Early Dementia (PrAISED), following changes required by the COVID-19 pandemic. BMJ open 2020; 10: e039305

[13] Nelson, J. Using conceptual depth criteria: addressing the challenge of reaching saturation in qualitative research. Qual Res 2017; 17: 554–70

[14] Braun, V, Clarke, V. Reflecting on reflexive thematic analysis. Qual Res Sport Exer Health 2019; 11: 589–97

[15] NVivo qualitative data analysis Software. Melbourne, Australia: QSR International Pty Ltd. 2018

[16] Burgui, D. Physical and mental wellbeing during the Covid-19 crisis. In: 30^th^ Alzheimer Europe Conference: Dementia in a changing world, 2020. Poster P01.37

[17] Rock, B, O’Philbin, L. Emergency Psychosocial Supports during Covid-19 for People with Dementia and their Families. In: 30^th^ Alzheimer Europe Conference: Dementia in a changing world, 2020. Presentation SS3.04

[18] Hickey, G, Brearley, S, Coldham et al. Guidance on co-producing a research project. Southampton: INVOLVE. 2018

[19] European Working Group of People with Dementia. My second new life: adapting after COVID-19. In: 30^th^ Alzheimer Europe Conference: Dementia in a changing world, 2020. Presentation SS1

[20] National Institute for Health and Clinical Excellence. Behaviour Change at Population, Community and Individual Levels. NICE Public Health Guidance. London: NICE; 2007.

[21] De Biase, S, Cook, L, Skelton, DA et al. The COVID-19 rehabilitation pandemic. Age and Ageing. 2020.

[22] Tsai, LL, McNamara, RJ, Dennis, SM, et al. Satisfaction and experience with a supervised home-based real-time videoconferencing telerehabilitation exercise program in people with chronic obstructive pulmonary disease (COPD). Int Jour Tel 2016; 8: 27

[23] Kettlewell, JL. Evaluating Brain-in-Hand for adults with an acquired brain injury. PhD. Thesis. University of Nottingham 2020.

[24] Van der Roest, H, Prins, M, Stolte, E, Steinmetz, S, Van Tilburg, T. Consequences of social isolation and uptake of communication technologies in Dutch nursing homes. In: 30^th^ Alzheimer Europe Conference: Dementia in a changing world, 2020. Poster P16.05

[25] Fabbo, A, Baglieri, A, Manni, B, Pellitta, A, Turci, M. The management of older people with Dementia and COVID-19 in nursing home. In: 30^th^ Alzheimer Europe Conference: Dementia in a changing world, 2020. Poster P24.03

[26] D’Anastasio, C, Ribani, V Barbani et al. Social media and virtual gathering among persons with dementia and their carers to face Covid-19 outbreak. In: 30^th^ Alzheimer Europe Conference: Dementia in a changing world, 2020. Poster P01.38

[27] Di Lorito, C, Duff, C, Rogers, C, Tuxworth, J et al. Tele-rehabilitation for people with dementia in the COVID-19 pandemic: A case-study. 2020. Submitted for publication.

[28] Vernooij-Dassen, M. Social distancing: protection and risk. In: 30^th^ Alzheimer Europe Conference: Dementia in a changing world, 2020. Poster P08.01

